# Urban infrastructure and spatiotemporal environmental features for *EGFR*-mutant lung cancer

**DOI:** 10.64898/2026.05.18.26353481

**Authors:** Danni Lu, Luchang Cui, Nick Kunz, Michelle Wong, Mohammad Tayarani, James P. Solomon, Christine A. Garcia, Nasser K. Altorki, Eunji Choi, H. Oliver Gao, Yiwey Shieh

**Affiliations:** Systems Engineering Program, Cornell University, Ithaca, NY 14853, USA; Department of Population Health Sciences, Weill Cornell Medicine, New York, NY 10065, USA; School of Civil and Environmental Engineering, Cornell University, Ithaca, NY 14853, USA; Department of Pathology and Laboratory Medicine, Weill Cornell Medicine, New York, NY 10065, USA; Department of Medicine, Division of Hematology and Oncology, Weill Cornell Medicine, New York, NY 10065, USA; Department of Cardiothoracic Surgery, Weill Cornell Medicine, New York, NY 10065, USA

**Keywords:** air pollution, *EGFR* mutations, machine learning, urban environment, lung cancer, environmental epidemiology

## Abstract

**Background:** Lung cancer in never-smokers is rising, with a substantial proportion harboring the *EGFR* mutation. While fine particulate matter (PM_2.5_) is a recognized risk factor, other intervenable pollutants and built environmental factors remain unknown.

**Objectives:** To identify urban characteristics associated with *EGFR*-mutant (vs. wild-type) lung cancer using high-resolution spatiotemporal data.

**Methods:** We analyzed 2,699 lung cancer patients with documented *EGFR* status treated at a high-volume academic medical center in New York City. Patient residential addresses were linked to high-resolution (300m×300m) 5-year cumulative exposures to 3 air pollutants and 26 urban features. We developed Light Gradient Boosting Machine (LightGBM) models to classify *EGFR* status, comparing a basic clinical model with established predictors (Asian, female, never-smoking status, and adenocarcinoma histology) to an extended model with additional urban factors. Predictive performance was assessed based on discrimination (AUC).

**Results:** We included 2,699 patients, of whom 54.1% were female and 25.8% self-identified as Asian, 11.2% as Black, and 7.4% as Hispanic; and 29% had *EGFR-*mutated cancer. The extended model showed modest improvements in discrimination (AUC: 0.775 [95% CI, 0.739-0.809] vs. 0.768 [0.723-0.811]), compared to the clinical model. Newly identified factors for *EGFR*-mutant status included black carbon (BC), nitrogen dioxide (NO_2_), proximity to airports, reduced access to public transportation, elevated noise levels, and lead exposure.

**Conclusions:** Traffic-related pollutants (BC, NO_2_) from diesel engines and motor vehicles, and proximity to airports, were among the novel spatiotemporal features associated with *EGFR*-mutant lung cancer. These results may inform policy interventions.

## Introduction

Globally, lung cancer (LC) retains its status as the predominant cause of cancer mortality and ranks third in incidence [1, 2]. While smoking is the primary risk factor, the epidemiological landscape of LC is shifting [3]. A concerning global trend is the rising absolute number of LC cases among never-smokers [4–6]. Patients with LC without history of smoking are molecularly unique, with *EGFR* mutations as the predominant oncogenic driver [4, 7]. *EGFR* mutation frequency differs across populations, occurring in approximately 47% of cases in the Asia-Pacific region, 15% in Europe, and 22% in North America [7–9]. This molecular subtype is also more common in women, younger patients, and those with adenocarcinoma histology [4, 5, 7].

Recent studies suggest that fine particulate matter (PM_2.5_) promotes tumorigenesis in *EGFR*-mutant LC, underscoring the importance of environmental factors in disease etiology [5, 10]. Given the complexity of ambient air pollution, other directly intervenable pollutants and urban environment factors also warrant investigation [11, 12]. A comprehensive understanding of *EGFR*-mutant LC risk, considering both individual pollutants and urban environmental factors, can enable targeted policy and public health interventions. Although a limited number of studies have examined built environment factors and LC risk [13–15], most have relied on population-level or area-level average exposures measured over relatively large geographic units, rather than individual-level exposures. Moreover, prior research has often overlooked localized urban features—such as housing quality, transit access, and industrial facilities—and has primarily employed linear modeling approaches that may fail to capture complex, high-dimensional relationships within urban environments [16]. Finally, data integration remains limited, with studies using either high-quality clinical data with coarse environmental estimates or high-resolution environmental data with aggregated health outcomes [17]. To bridge this gap, studies leveraging high-resolution remote sensing and machine learning (ML) applied to clinically annotated hospital data are needed to enable simultaneous evaluation of detailed individual- and area-level factors at a scale previously infeasible [18].

We aimed to identify environmental drivers of *EGFR*-mutant (vs. wild-type) LC by integrating detailed clinical data with high-resolution spatiotemporal environment features using advanced ML. These methods can capture complex nonlinear relationships and high-order interactions among predictors, particularly in high-dimensional settings where traditional regression models may be limited [18]. We also aimed to enhance interpretability by quantifying the contributions of identified factors, potentially revealing new targets for public health interventions and prevention strategies for the growing population of never-smoking *EGFR*-mutant lung cancer patients.

## Methods

### Study Population

Data were obtained from the Meyer Cancer Center Molecularly Enhanced Lung Cancer Database (MCC-MELD), an electronic health record-based registry comprising 9,573 LC cases from a large, diverse catchment area in New York City [19]. The database includes three NewYork-Presbyterian (NYP)-affiliated clinical sites: Weill Cornell Medicine in Manhattan, NYP Queens, and NYP Brooklyn Methodist Hospital. MCC-MELD integrates detailed clinical information and genomic testing results, leveraging natural language processing (NLP) to extract additional key data from free-text clinical notes. Tumor sequencing data and NLP algorithms are combined to accurately capture *EGFR* mutation status [19]. Ethical oversight of this work was provided by the Weill Cornell Medicine Institutional Review Board (Protocols 24-04027328 and 24-05027508).

Our analysis included patients residing in New York City (n = 7,479) with known smoking status (n = 5,679) and tumors with available histology (n = 5,425) and confirmed *EGFR* mutation status (n = 2,699) (**Supplementary Figure 1**).

### Feature engineering for air pollution and urban factors

We generated a 500-m buffer around each patient’s residential address ascertained at the time of diagnosis to characterize local environmental exposure, following methods widely applied in environmental epidemiology and health-geography studies [20–22]. Historical air pollution data were integrated, focusing on 5-year cumulative exposures to PM_2.5_, black carbon (BC), and nitrogen dioxide (NO_2_), obtained from the NYCCAS air pollution raster dataset. Each patient’s 500-m buffer was linked to a 300 m × 300 m air pollution grid to calculate mean pollutant concentrations from 2015 to 2023, with cumulative exposures computed for the five years preceding diagnosis [5, 23, 24].

Transportation-related exposure and infrastructure metrics included annual average daily traffic (AADT), vehicle miles traveled (VMT), truck routes, 5-miles airports buffer [25, 26], bike routes, bus depots and terminals, bus lanes, bus stops, subway lines and stations, as well as traffic-related deaths, serious injuries, and traffic noise. Stationary emission sources encompassed regulated industrial facilities, chemically intensive small businesses, and hazardous waste and solid-waste management facilities. Housing-related indicators, such as air-conditioning malfunctions, maintenance deficiencies, and lead service lines, were also incorporated. All transportation, emission, and housing data were obtained from the EJNYC Mapping Tool website.

To capture land-use mix information, we integrated two sources: OpenStreetMap (OMS) land-use layers downloaded via python using “osmnx” package [27], and official land-use data from the Primary Land Use Tax Lot Output (PLUTO) database [28]. These were combined to provide more complete coverage, with additional metrics including the ratio of built area to green space from EJNYC Mapping Tool website.

A consistent buffer-based approach was applied across all datasets to derive traffic-related, housing, and land-use metrics [29, 30]. For linear features (e.g., roads, transit lines), total lengths or counts within each buffer were calculated; for point features (e.g., bus stops, subway stations), total counts per buffer were determined; and for polygon-based variables (e.g., traffic-related mortality, areas exposed to noise > 45 dB), mean values within each buffer were computed (**Supplementary Table 1**). Spatial data processing and feature extraction, specifically using the Spatial Join function, were conducted using ArcGIS Pro. All statistical analyses and machine learning model development were performed using Python (version 3.12) with the autogluon (v 1.3.1), lightgbm (v 4.6.0), scikit-learn (v 1.4.2), and shap (v 0.46.0) libraries.

### Machine Learning (ML) Method

*Overview*. We employed a two-stage ML framework combining Automated Machine Learning (AutoML) with expert-guided refinement. First, the data were split into training and testing subsets using a stratified 70/30 split to preserve class proportions. AutoGluon trained, evaluated, and ensembled diverse models in the training set through automated hyperparameter optimization, bagging, and stacking [31, 32], improving generalizability and minimizing subjective tuning [33, 34]. LightGBM emerged as the top-performing model from AutoGluon’s leaderboard and was selected for further refinement based on empirical performance [35]. Additionally, bootstrap resampling (1,000 iterations) was used as an internal validation procedure to estimate the sampling distribution of performance metrics and derive their 95% confidence intervals. Detailed descriptions of the LightGBM algorithm, training process, and validation procedures are provided in **Supplementary Methods 1**.

#### Learning task

The primary learning task was to identify *EGFR*-mutant LC (vs. *EGFR* wild-type) patients.

#### Model Evaluation

Model performance was primarily assessed using the area under the receiver operating characteristic curve (ROC-AUC), which served as the main criterion for model comparison and selection. To convert predicted probabilities into binary classifications, the probability threshold was subsequently optimized to maximize the F1-score based on the precision-recall curve. Macro-averaged and weighted F1-scores were included to account for class imbalance and to evaluate performance across classes with equal and prevalence-weighted importance, respectively. These metrics were calculated on the held-out test set.

#### Model interpretability using SHAP

To improve transparency and interpretability of the final model, Shapley Additive exPlanations (SHAP) was applied to the Light GBM model [36]. SHAP uses a game-theoretic framework to assign feature contributions to individual predictions [37]. We visualized these contributions in a SHAP summary plot, showing both feature importance and how feature values influence prediction direction. Positive SHAP values indicate features that push the model toward *EGFR*-mutant status, whereas negative values push it away. Larger absolute SHAP values indicate stronger contributions to the prediction.

#### Sensitivity analysis

First, given the clinical relevance of *EGFR*-mutant LC among never-smokers, we repeated ML modeling in this subgroup. Second, because MCC-MELD includes three hospital sites (Weill Cornell Medicine, NYP Queens, NYP Brooklyn Methodist Hospital) with differing patient composition, catchment area characteristics, and data completeness, we conducted a subset analysis using the largest site (Weill Cornell Medicine) to ensure robust estimates. Finally, since *EGFR* testing was first approved in 2003 and widely adopted after 2019, many patients have unknown *EGFR* status. We therefore performed an additional ML analysis including patients with unknown *EGFR* status, treated as a three-class outcome (mutant, wild-type, unknown).

## Results

Among the 2,699 patients in the study cohort (**Table 1**), *EGFR*-mutant LC exhibited stronger female predominance (66.7% vs 49.0%) and earlier age of onset (68.6 vs. 69.6 years) compared with wild-type. Over half of patients with *EGFR*-mutant LC were never-smokers (51.1% vs. 15.2%), adenocarcinoma was the predominant histology (92.0%), and Asians represented the largest racial group (47.9%).

**Table 1.**
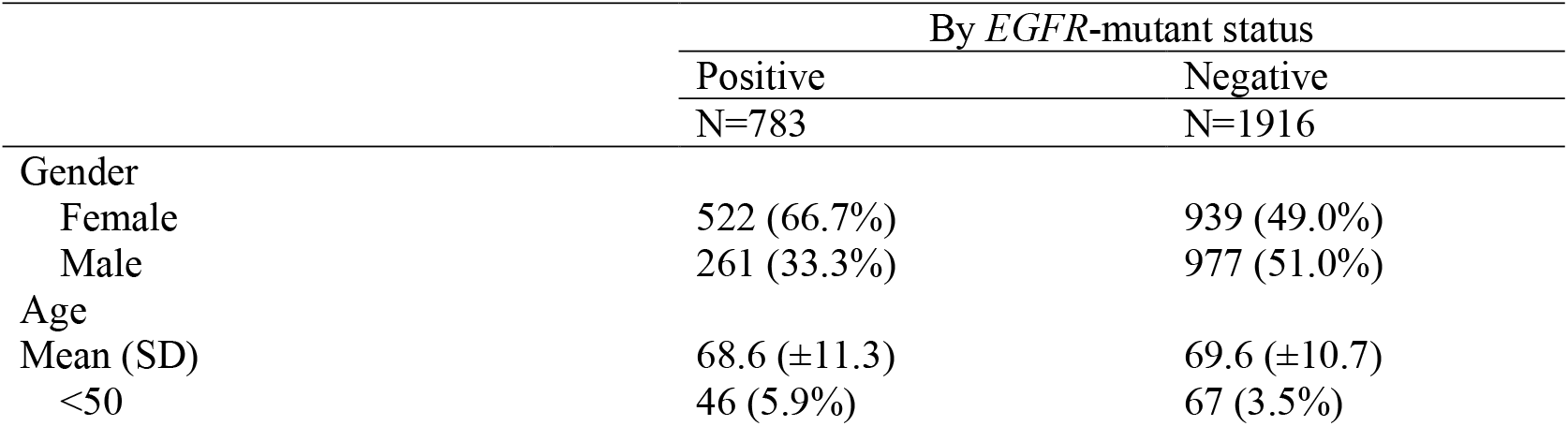
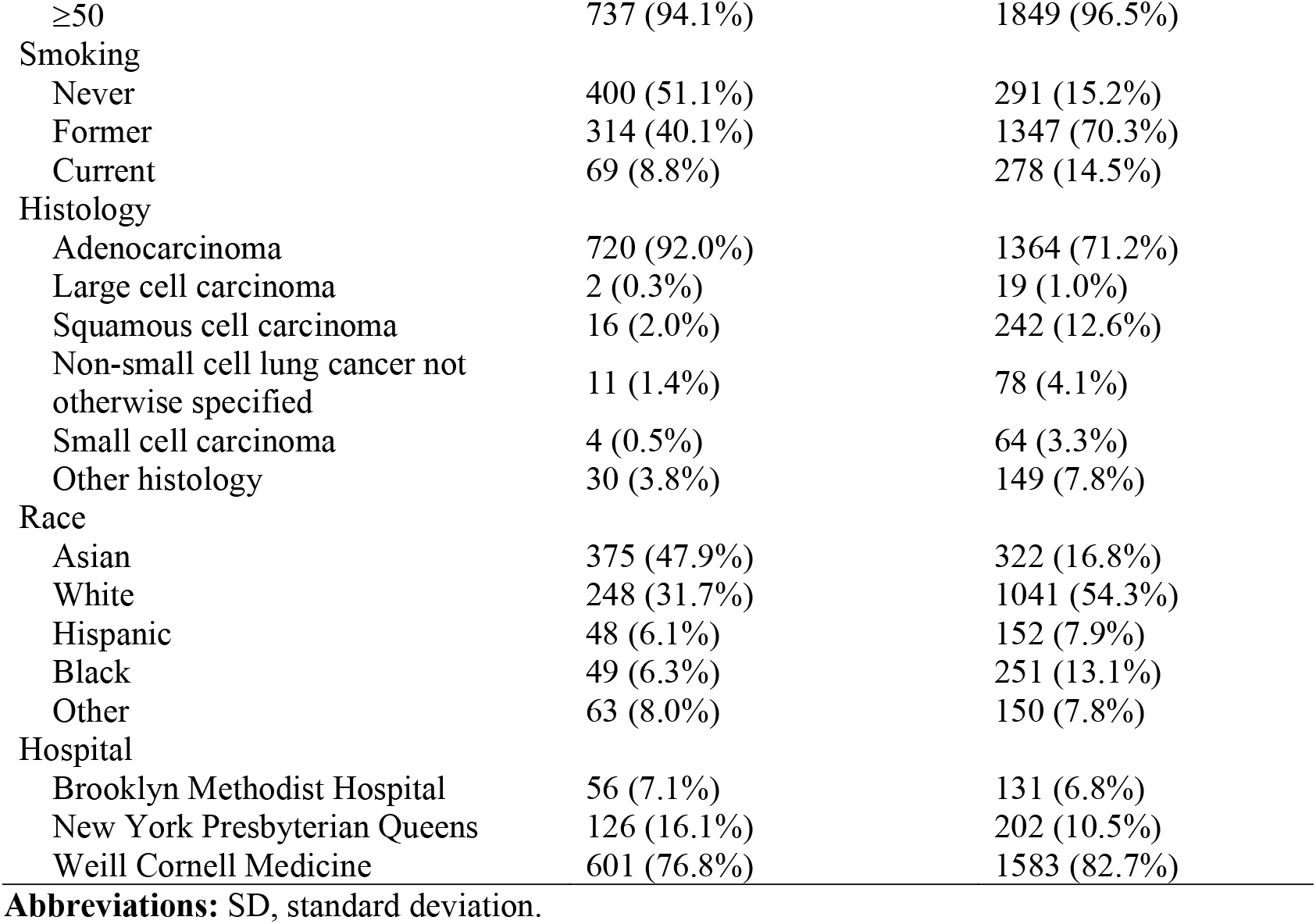
Patient-level Characteristics by *EGFR*-mutant LC Status.

The geographic distribution of LC cases varied by sex and *EGFR* status (**Figure 1**). Importantly, these patterns reflect where patients sought care within the participating hospital systems rather than actual geographic clustering or population-level disease prevalence. Among *EGFR*-mutant cases, female patients were more commonly located in Flushing, Queens, whereas male patients more often resided in Chinatown, Manhattan. Similar distributions were observed for *EGFR*-mutant versus wild-type cases by smoking status (never vs ever; **Supplementary Figure 2**), race/ethnicity (**Supplementary Figure 3**), and LC histology (**Supplementary Figure 4**).

**Figure 1.**
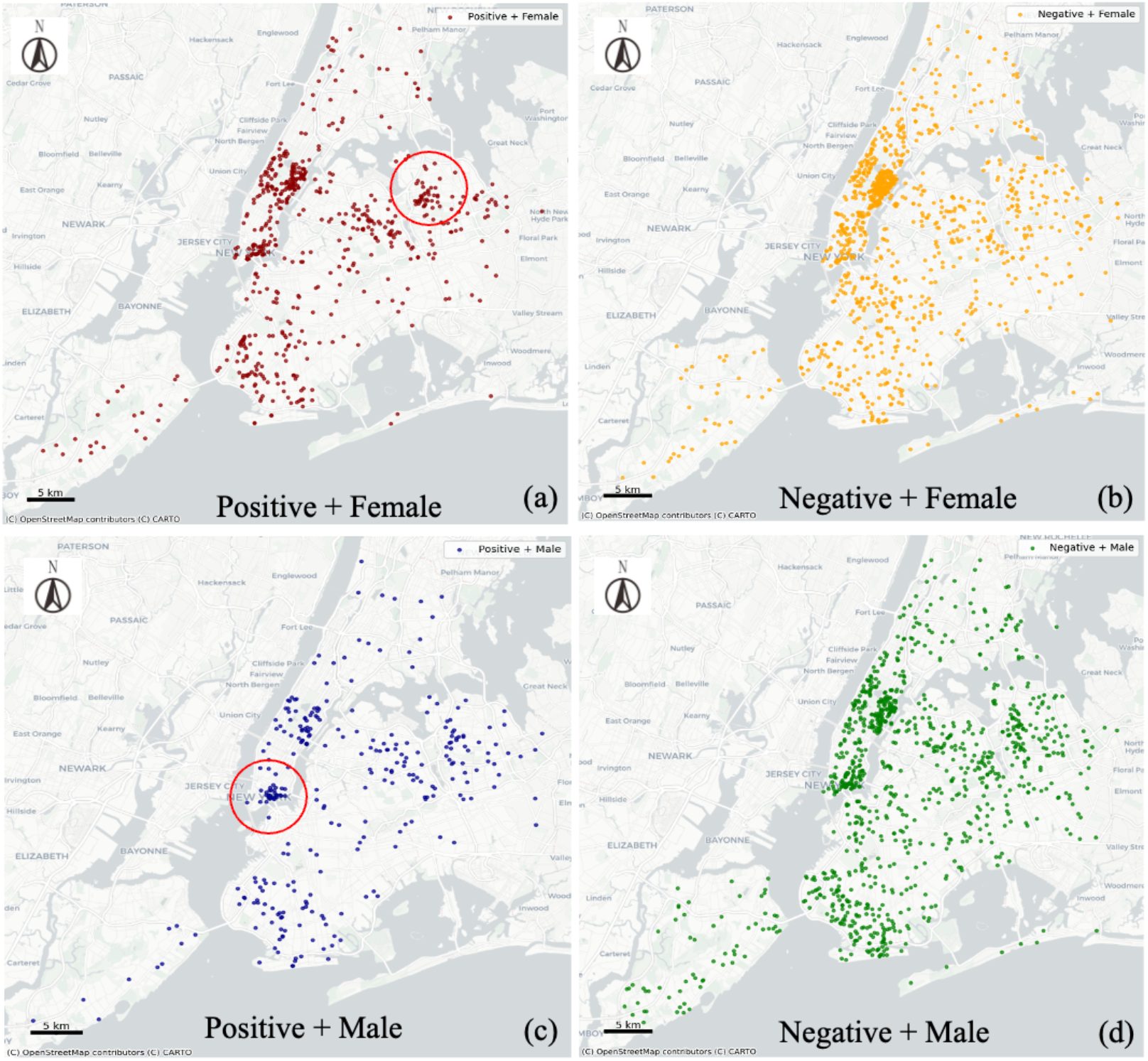
Distribution of Lung Cancer Cases by Sex and *EGFR*-mutant Status Across Meyer Cancer Center Catchment Areas at WCM (Manhattan), BMH (Brooklyn), and NYPQ (Queens). (a) *EGFR*-positive females, (b) *EGFR*-negative females, (c) *EGFR*-positive males, and (d) *EGFR*-negative males. **Note:** Each point represents a patient residential location. Red circles indicate areas with visually apparent concentrations of cases and are provided for illustrative purposes only.

Most environmental and transportation indicators were broadly similar between groups stratified by *EGFR* status (**Table 2**). However, several variables showed modest numerical differences. Patients with *EGFR*-mutant lung cancer resided in neighborhoods with slightly higher mean cumulative exposure to NO_2_ and lower levels of transit and active-transport infrastructure, including shorter mean bike-lane length and less bus and subway infrastructure (e.g., shorter bus route length and bus lane length, and shorter subway length). These neighborhoods also showed numerically higher indicators of traffic intensity, including higher mean vehicle miles traveled (VMT) and a greater proportion of residences located within 5 miles of an airport. Moreover, neighborhoods in which patients with *EGFR*-mutant lung cancers resided showed a numerically higher values for impervious built-area-to-green-space ratio, a numerically higher housing maintenance issue rate, and a higher percentage of chemically intensive business.

**Table 2.**
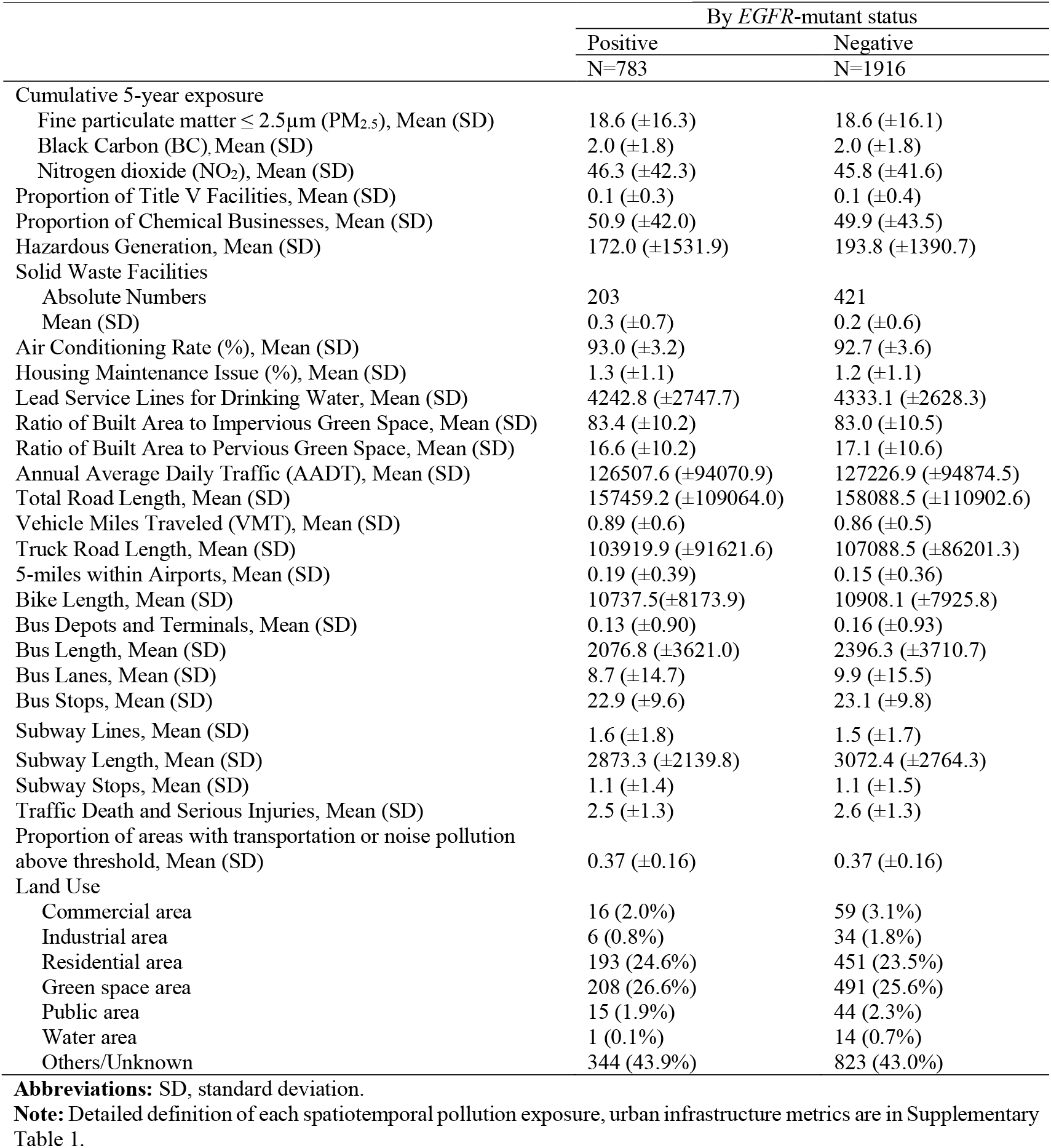
Built Environment Characteristics by *EGFR*-mutant LC status.

|  | By *EGFR*-mutant status |  |
| --- | --- | --- |
|  | Positive<br>N=783 | Negative<br>N=1916 |
| Cumulative 5-year exposure |  |  |
| Fine particulate matter $\leq 2.5\mu\text{m}$ (PM<sub>2.5</sub>), Mean (SD) | 18.6 ($\pm 16.3$) | 18.6 ($\pm 16.1$) |
| Black Carbon (BC), Mean (SD) | 2.0 ($\pm 1.8$) | 2.0 ($\pm 1.8$) |
| Nitrogen dioxide (NO<sub>2</sub>), Mean (SD) | 46.3 ($\pm 42.3$) | 45.8 ($\pm 41.6$) |
| Proportion of Title V Facilities, Mean (SD) | 0.1 ($\pm 0.3$) | 0.1 ($\pm 0.4$) |
| Proportion of Chemical Businesses, Mean (SD) | 50.9 ($\pm 42.0$) | 49.9 ($\pm 43.5$) |
| Hazardous Generation, Mean (SD) | 172.0 ($\pm 1531.9$) | 193.8 ($\pm 1390.7$) |
| Solid Waste Facilities |  |  |
| Absolute Numbers | 203 | 421 |
| Mean (SD) | 0.3 ($\pm 0.7$) | 0.2 ($\pm 0.6$) |
| Air Conditioning Rate (%), Mean (SD) | 93.0 ($\pm 3.2$) | 92.7 ($\pm 3.6$) |
| Housing Maintenance Issue (%), Mean (SD) | 1.3 ($\pm 1.1$) | 1.2 ($\pm 1.1$) |
| Lead Service Lines for Drinking Water, Mean (SD) | 4242.8 ($\pm 2747.7$) | 4333.1 ($\pm 2628.3$) |
| Ratio of Built Area to Impervious Green Space, Mean (SD) | 83.4 ($\pm 10.2$) | 83.0 ($\pm 10.5$) |
| Ratio of Built Area to Pervious Green Space, Mean (SD) | 16.6 ($\pm 10.2$) | 17.1 ($\pm 10.6$) |
| Annual Average Daily Traffic (AADT), Mean (SD) | 126507.6 ($\pm 94070.9$) | 127226.9 ($\pm 94874.5$) |
| Total Road Length, Mean (SD) | 157459.2 ($\pm 109064.0$) | 158088.5 ($\pm 110902.6$) |
| Vehicle Miles Traveled (VMT), Mean (SD) | 0.89 ($\pm 0.6$) | 0.86 ($\pm 0.5$) |
| Truck Road Length, Mean (SD) | 103919.9 ($\pm 91621.6$) | 107088.5 ($\pm 86201.3$) |
| 5-miles within Airports, Mean (SD) | 0.19 ($\pm 0.39$) | 0.15 ($\pm 0.36$) |
| Bike Length, Mean (SD) | 10737.5 ($\pm 8173.9$) | 10908.1 ($\pm 7925.8$) |
| Bus Depots and Terminals, Mean (SD) | 0.13 ($\pm 0.90$) | 0.16 ($\pm 0.93$) |
| Bus Length, Mean (SD) | 2076.8 ($\pm 3621.0$) | 2396.3 ($\pm 3710.7$) |
| Bus Lanes, Mean (SD) | 8.7 ($\pm 14.7$) | 9.9 ($\pm 15.5$) |
| Bus Stops, Mean (SD) | 22.9 ($\pm 9.6$) | 23.1 ($\pm 9.8$) |
| Subway Lines, Mean (SD) | 1.6 ($\pm 1.8$) | 1.5 ($\pm 1.7$) |
| Subway Length, Mean (SD) | 2873.3 ($\pm 2139.8$) | 3072.4 ($\pm 2764.3$) |
| Subway Stops, Mean (SD) | 1.1 ($\pm 1.4$) | 1.1 ($\pm 1.5$) |
| Traffic Death and Serious Injuries, Mean (SD) | 2.5 ($\pm 1.3$) | 2.6 ($\pm 1.3$) |
| Proportion of areas with transportation or noise pollution above threshold, Mean (SD) | 0.37 ($\pm 0.16$) | 0.37 ($\pm 0.16$) |
| Land Use |  |  |
| Commercial area | 16 (2.0%) | 59 (3.1%) |
| Industrial area | 6 (0.8%) | 34 (1.8%) |
| Residential area | 193 (24.6%) | 451 (23.5%) |
| Green space area | 208 (26.6%) | 491 (25.6%) |
| Public area | 15 (1.9%) | 44 (2.3%) |
| Water area | 1 (0.1%) | 14 (0.7%) |
| Others/Unknown | 344 (43.9%) | 823 (43.0%) |

**Abbreviations:** SD, standard deviation.
**Note:** Detailed definition of each spatiotemporal pollution exposure, urban infrastructure metrics are in Supplementary Table 1.

The final LightGBM model displayed the strongest performance for identifying *EGFR*-mutant lung cancer (**Figure 2**). Key features associated with *EGFR*-mutant LC included never-smoking status, Asian race, adenocarcinoma histology, and female sex. In addition, the model identified environmental contributors, including neighborhood noise levels, cumulative exposures to NO_2_ and BC, lead exposure, and proximity to airports. The SHAP summary plot (**Figure 2b**) shows the relative importance of these features and the direction of their contributions within the model. Never-smoking, Asian race, adenocarcinoma histology, and female sex showed predominantly positive contributions. Environmentally, higher noise levels, higher cumulative exposure to NO_2_ and BC, higher lead exposure, and living within 5 miles of an airport associated with *EGFR*-mutant lung cancer. Shorter subway length and shorter bus length also contributed to the positive direction, indicating that lower transit infrastructure is associated with *EGFR*-mutant status. In an extended model that integrated clinical with additional air pollution and urban features, LightGBM achieved the highest overall performance (accuracy = 0.761 [95% CI, 0.731-0.790]; AUC = 0.775 [95% CI, 0.739-0.809]) (**Table 3**). Adding built-environment features produced a modest but consistent performance gain relative to patient-only models. For LightGBM, the category-specific F1 score for the *EGFR*-mutant positive class increased from 0.586 [95% CI, 0.526-0.648] using patient features only to 0.625 [95% CI, 0.574-0.674] when environmental variables were added (**Table 3**).

**Figure 2.**
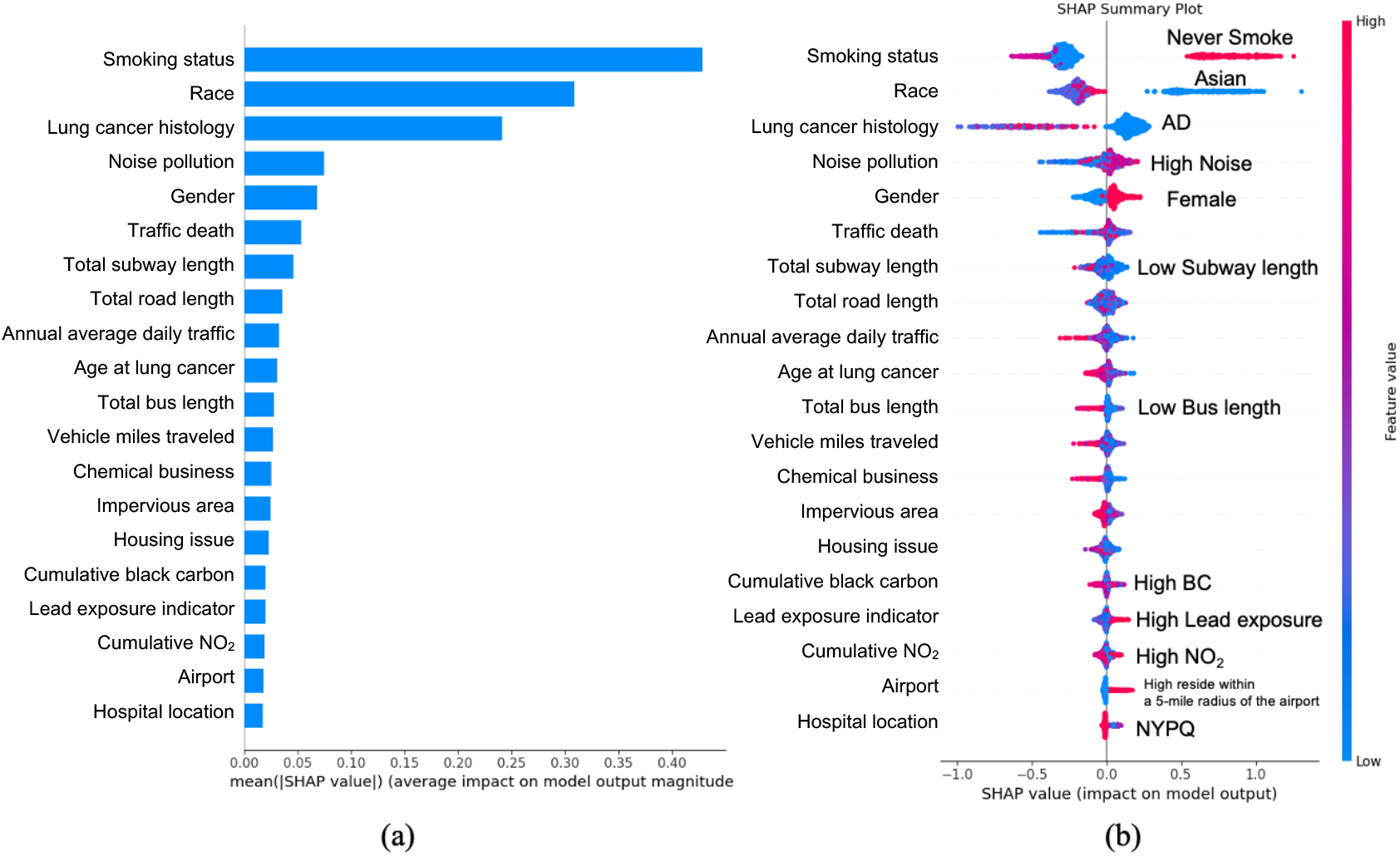
Feature Importance of Predictors Associated with *EGFR*-mutant Lung Cancer Using Light Gradient Boosting Machine.Panel (a) illustrates the global feature importance, ranking predictors by their mean absolute SHAP values to indicate the magnitude of their overall contribution to the model. Panel (b) presents the SHAP summary plot, visualizing the direction and distribution of feature effects. Each dot represents a single observation. The color gradient indicates the feature value (pink for high values, blue for low values). Regarding the x-axis, a positive SHAP value indicates that the feature increases the predicted probability of *EGFR*-mutant lung cancer, while a negative SHAP value indicates a decrease in probability. **Abbreviations:** *EGFR*, epidermal growth factor receptor; SHAP, Shapley Additive exPlanations; AD, adenocarcinoma; Noise pollution, transportation and noise pollution above threshold; Airport, resides within 5 miles of an airport. New

**Table 3.**
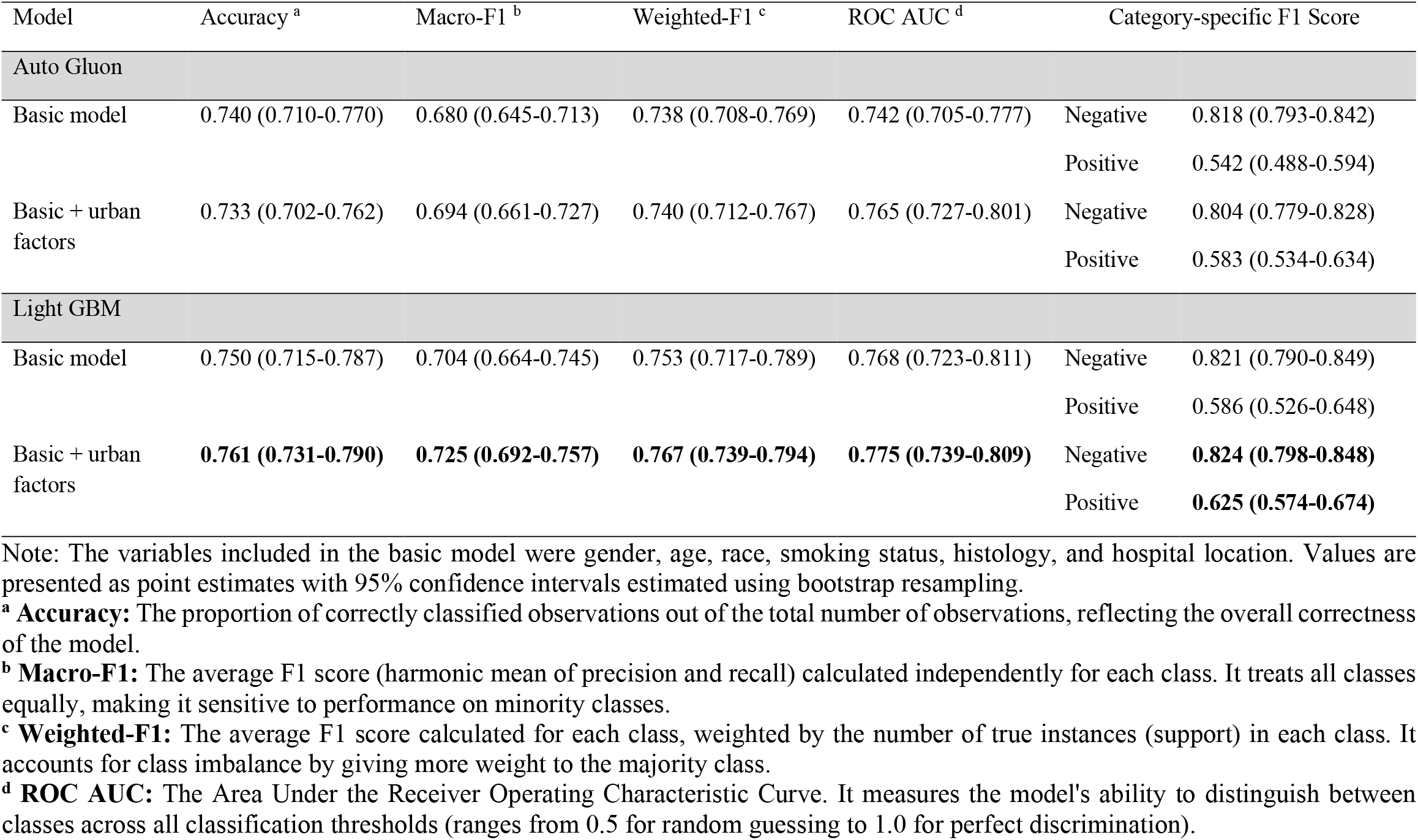
Binary Classification Performance.

Sensitivity ML analyses consistently identified key associated factors. Additionally, the model restricted to never-smokers identified traffic intensity, reflected by higher vehicle miles traveled (VMT) and annual average daily traffic (AADT) (**Supplementary Table 2; Supplementary Figure 5**). A model based solely on data from the Weill Cornell Medicine site included many of the same variables as our main model but identified additional site-specific urban infrastructure features, such as poor housing air-conditioning conditions and high number of bus lanes (**Supplementary Table 3; Supplementary Figure 6**). Multiclass model performance is summarized in **Supplementary Table 4**; beyond the clinical predictors driving the binary model, the multiclass framework highlighted additional environmental contributors, particularly longterm PM_2.5_ exposure, which helped distinguish between the three outcome categories (**Supplementary Figure 7**).

## Discussion

We built a ML model integrating high-resolution spatiotemporal features with individual-level clinical data to examine the associations between clinical and environmental features with *EGFR*-mutant versus wildtype LC. Within this multivariable framework, we redemonstrated established, positive associations of never-smoking status, self-reported Asian race, female sex, and adenocarcinoma histology with *EGFR*-mutant LC. Moreover, we identified novel environmental features preferentially associated with *EGFR*-mutant LC, including cumulative 5-year exposure to BC and NO_2_, proximity to airports, noise pollution, and lead exposure. These features provided robust predictive signals that were independent of, and complementary to, clinical variables. Our study demonstrates that ML frameworks can identify and characterize learnable environmental patterns associated with *EGFR*-mutant LC, establishing a critical link between the urban built environment and molecular epidemiology.

Our results highlight the potentially complex, non-linear interactions between demographic, clinical, behavioral, and environmental factors and tumor *EGFR* status. We found modest group-level differences in patient-level and built environment characteristics by *EGFR* mutation status and the lack of strong contrasts across most variables (**Tables 1** and **2**). In this setting, supervised machine-learning models may be able to identify individual-level patterns and multivariable relationships that may not be detectable through traditional descriptive analyses. Indeed, the incorporation of built environment factors into the LightGBM framework resulted in a modest increase in the overall ROC-AUC. Most notably, the model demonstrated enhanced predictive F1-score specifically for the *EGFR*-mutant-positive class. This performance boost confirms that the residential environment constitutes an extrinsic determinant of LC subtype, complementary to clinical factors [38]. In addition, the reproducibility of established factors with *EGFR*-mutant LC supports the predictive validity of our modeling approach.

Our model identified several previously unrecognized air pollutants and built environmental factors associated with *EGFR* positivity, supporting the potential influence of multi-pollutant exposures and pollution-inducing factors. Specifically, we observed significant associations between *EGFR*-mutant LC status and high noise exposure, cumulative 5-year exposure to black carbon (BC), cumulative 5-year exposure to NO_2_, and lead. Both NO_2_ and BC primarily impact respiratory physiology. NO_2_ is a well-established risk factor for LC, with meta-analyses showing that every 10 μg/m^3^ increment in exposure is associated with a 10% higher risk [39–41]. Similarly, BC exposure also impairs lung function, particularly in former smokers, suggesting heightened vulnerability in individuals with prior airway damage [42]. Although prior studies have established epidemiologic and mechanistic evidence linking long-term PM_2.5_ exposure to the development of *EGFR*-mutant LC, PM_2.5_ did not emerge as an independent predictor in the final model. This result should be interpreted within a multi-pollutant framework, as PM_2.5_ shares substantial source overlap with traffic-related pollutants such as NO_2_ and BC in urban environments. Consequently, the inclusion of correlated pollutants may attenuate individual effect estimates without undermining the established role of PM_2.5_ in lung carcinogenesis.

Furthermore, residential proximity to major transportation hubs defined using a 5-mile buffer of airports based on prior literature [25, 26] was identified as a model-informative feature, likely because it captures geographic proximity to airport related emission sources rather than representing an independent environmental exposure effect. Residential proximity to airports represents a unique convergence of these physical and chemical risks [43]. Residents near airports face a dual burden: chronic exposure to high-decibel aviation noise and inhalation of complex combustion byproducts. These emissions, which include NO_x_, carbon monoxide, and specific carcinogens derived from jet fuel, create a distinct toxicological profile compared to road traffic [44]. While some studies suggest that airport-related ultrafine particles are more strongly associated with squamous cell carcinoma rather than adenocarcinoma [43], our model’s identification of airport proximity as a predictor for *EGFR*-mutant LC suggests that other components, such as fine particulate matter [45] or volatile organic compounds from unburned fuel, may be related to *EGFR* positivity. The persistence of jet fuel odors in summer months further highlights the potential for significant exposure to organic solvents [43], which may act synergistically with noise-induced stress to promote tumor growth.

Our findings offer hypotheses regarding urban environmental contributors to *EGFR*-mutant LC risk that, if validated, may inform future policy and public health interventions. Our finding that BC, NO_2_, noise pollution, and lead exposure may be informative for *EGFR* positivity are particularly relevant in the context of New York City’s evolving environmental policies, such as recent emission control mandates and congestion mitigation strategies [46]. Further elucidation of the link between specific pollutants and LC subtypes may enable policies that reduce local BC and NO_2_ levels, which could in turn reduce the incidence of *EGFR*-mutant LC. Furthermore, confirmation of airport proximity and noise as distinct risk factors would support stricter zoning regulations and noise abatement programs in residential areas bordering major transportation hubs [47]. On a public health level, the environmental signatures identified in this study, if validated, may help identify neighborhoods where combined environmental exposures are associated with a higher likelihood of *EGFR*-mutant LC. Such information could support targeted educational outreach, environmental monitoring, and community-level disease surveillance in area with elevated environmental risk profiles [48].

Our study presents several key strengths that advance the understanding of built environmental determinants in LC. First, we achieved an innovative integration of high-resolution urban infrastructure and environmental data with detailed individual-level clinical characteristics using GIS-based spatial linkage within patient-centered buffer zones. This multi-modal approach enables the discovery of risk factors beyond traditional epidemiology and allows for the evaluation of their joint effects on disease pathogenesis. Second, the study specifically stratifies patients by *EGFR*-mutant status. By focusing on this distinct subtype, we provide critical insights into the impact of the built environment on *EGFR*-mutant LC, extending our understanding beyond patient characteristics alone. Third, the application of advanced machine learning techniques (LightGBM) coupled with SHAP interpretability offers a dual advantage: it captures complex, non-linear interactions to ensure robust predictive performance while maintaining model transparency to visualize the contribution of each feature. Finally, our findings have clear public health relevance, providing an evidence base for targeted interventions aimed at mitigating environmental contributors to LC risk.

Despite these strengths, several limitations should be acknowledged. First, as an observational, case-only study, our findings reflect statistical associations rather than causal relationships. The absence of a cancer-free comparison group limits inference regarding disease risk, and unmeasured confounding factors, such as lifestyle behaviors (e.g., diet and physical activity) or indoor air pollution, could not be fully considered [49]. Second, regarding exposure assessment, our reliance on the residential address at the time of diagnosis to estimate five-year cumulative exposure introduces potential exposure misclassification. This method does not account for residential mobility (migration) during the latency period, nor does it capture occupational exposures or time-activity patterns (i.e., time spent outside the home), which may affect actual dosage [50, 51]. Third, the missingness of *EGFR*-mutant LC status in a subset of the cohort may introduce selection bias, as testing availability often correlates with socioeconomic status or clinical presentation [19, 52]. In addition, the cohort may be subject to broader institutional selection biases. The proportion of adenocarcinoma in our cohort appeared higher than that reported in some other populations, which may reflect referral patterns, differences in clinical management, or variation in tumor selection for sequencing across hospitals and over time, particularly before reflex sequencing was fully established. As such, the representativeness and generalizability of our findings may be limited [53]. Furthermore, although the use of SHAP values allows us to capture complex non-linear patterns that linear models often miss, it does not generate a fixed global effect estimate like an odds ratio [36]. Unlike logistic regression, which provides a single coefficient for the entire population, our model calculates contributions at the individual level. Consequently, this methodological difference limits the feasibility of direct quantitative comparisons with existing epidemiological literature relying on linear effect estimates [54]. Finally, our findings have not yet been independently validated in external datasets. Future studies should aim to validate these findings in independent cohorts and across diverse environmental contexts.

## Conclusions

Our study demonstrates that machine learning frameworks can identify and characterize learnable environmental patterns associated with *EGFR*-mutant LC, establishing a critical link between the urban built environment and molecular oncology. Our approach validates the feasibility of modeling the complex, non-linear joint effects of individual characteristics and environmental exposures using real-world data, effectively broadening the scope of disease prediction beyond patient demographics. Crucially, the identification of these distinct spatial patterns suggests that specific, modifiable environmental factors such as cumulative BC, NO_2_, and noise exposure may be associated with *EGFR*-mutant LC status in populations historically deemed less susceptible, such as non-smokers. Consequently, our findings advocate for a two-pronged translation strategy: the integration of learned environmental signatures when considering predilection for lung cancer subtype, and the implementation of targeted urban health interventions to mitigate exposure to potential carcinogens.

## Supporting information

Supplementary Table 1-4, Supplementary Figure 1-7, Supplementary Methods 1, and will be used for the link to the file on the preprint site.

## Data Availability

The patient-level clinical and genomic data used in this study are not publicly available due to patient privacy protections, institutional policies, and institutional review board restrictions. Publicly available environmental and urban infrastructure datasets used in this study are available from their original sources, as described in the Methods section. Aggregate results supporting the findings are provided in the manuscript and supplementary materials.

## Acknowledgements

This study was supported by a pilot grant from the Weill Cornell Medicine Meyer Cancer Center EGFR-mutant Lung Cancer Initiative and a Ritu Banga Healthcare Disparities Research Award to Y. Shieh.

