## Supplementary Table 1-4, Supplementary Figure 1-7, Supplementary Methods 1, and will be used for the link to the file on the preprint site. for "Urban infrastructure and spatiotemporal environmental features for *EGFR*-mutant lung cancer"

### Supplementary Files

**Supplementary Table 1.** Spatiotemporal Pollution Exposure, Urban Infrastructure Metrics
**Supplementary Table 2.** Binary Classification Performance Using Never Smoker Patients
**Supplementary Table 3**. Binary Classification Performance Using WCM Data Only
**Supplementary Table 4**. Multi-Classification Performance

**Supplementary Figure 1.** Cohort Selection Diagram
**Supplementary Figure 2.** Distribution of Lung Cancer Cases by Never or Ever Smoking Status and *EGFR*-mutant Status Across Meyer Cancer Center Catchment Areas at WCM (Manhattan), BMH (Brooklyn), and NYPQ (Queens).
**Supplementary Figure 3**. Distribution of Lung Cancer Cases by Race and *EGFR*-mutant Status Across Meyer Cancer Center Catchment Areas at WCM (Manhattan), BMH (Brooklyn), and NYPQ (Queens).
**Supplementary Figure 4**. Distribution of Lung Cancer Cases by Histology and *EGFR*-mutant Status Across Meyer Cancer Center Catchment Areas at WCM (Manhattan), BMH (Brooklyn), and NYPQ (Queens).
**Supplementary Figure 5.** Feature Importance of Model Using Never Smoker Patients
**Supplementary Figure 6.** Feature Importance of Model Using WCM Data Only
**Supplementary Figure 7.** Feature Importance of Predictors Associated with *EGFR*-mutant Lung Cancer Using Light Gradient Boosting Machine (LightGBM)

**Supplementary Methods 1.** Light Gradient Boosting Model (LightGBM)

**Supplementary Table 1.** Spatiotemporal Pollution Exposure, Urban Infrastructure Metrics (Emission, House, Vehicle Operation, Transportation Infrastructures and Land Use)

| Category | Definition | Integration |
|---|---|---|
| **Dependent Variable** | | |
| EGRF Mutated Lung Cancer | EGFRm LC vs other LC: Negative=0, Positive=1, Unknown=2 | Point Buffer Analysis |
| **Independent Variables** | | |
| **Patients** | | |
| Race | Self-reported Race Categories: Asian=1, While=2, Hispanic=3, Black=4, Other=5 | Individual Attribute |
| Gender | Biological Sex of the Patient: Male=1, Female=2 | |
| Age | Age in years at lung cancer diagnosis. | |
| Smoke Status | Smoking history: Former=1, Unknown=2, Current=3, Never=4 | |
| Histology | Histological subtype of lung cancer, e.g., Adenocarcinoma (AD=1), Squamous (SQ=2), NSCLC NOS=3, Small Cell (SC=4), Other (OTH=5), Large Cell (LC=6), Missing/Unknown (NA=0). | |
| **Historical Air Pollution** | | |
| Cumulative $PM_{2.5}$, BC, $NO_2$ | Average and cumulative $PM_{2.5}$, BC, $NO_2$ (Nitrogen Dioxide) during the 5-year periods prior to the diagnosis year | Point Buffer Temporal Cumulative |
| **Emission** | | |
| Title V Facilities | Count of federally regulated industrial facilities with permitted air emissions (e.g., VOCs, $NO_x$, CO, $PM_{2.5}$, $SO_2$, HAPs) within buffer. | Buffer Count |
| Chemically Intensive Small Business | Establishments with intensive chemical usage (e.g., automotive repair/body shops, dry cleaning). | Buffer Count |
| Hazardous Waste Material Storage | Facilities permitted to store or manage hazardous waste. | Buffer Count |
| Solid Waste Management Facilities | Establishments with intensive chemical usage (e.g., automotive repair/body shops, dry cleaning). | Buffer Count |
| **House** | | |
| Air Conditioning access | Percentage of households reporting functioning air conditioning. | Buffer Mean |
| Housing Maintenance Issues | Estimated number of households in the area reporting three or more maintenance deficiencies. | Buffer Mean |
| Lead Service Lines for Drinking Water | Population-normalized number of lead service lines per census tract. | Buffer Mean |
| Ratio of Built Area to Green Space | Percent impervious area and pervious (green) area for each community district; used to derive built/green ratio. | Buffer Mean |
| **Vehicle Operation and Transportation Infrastructures** | | |
| AADT | Annual Average Daily Traffic; total road length; VMT is length-weighted AADT which defined as AADT divided by total road length within buffer. | Line Sum and Ratio |
| Truck Route | Annual Average Daily Traffic; total road length; length-weighted AADT defined as AADT divided by total road length within buffer. | Line Sum |
| Airports | Five miles buffer of airports. | Buffer Count |
| Bike Lines and Routes | Length of dedicated bicycle lanes and marked cycling routes. | Line sum |
| Bus Depots and Terminals | Locations of bus depots/terminals (e.g., school, MTA, Port Authority). | Buffer Count |
| Bus Lanes and Bus Ways | Infrastructure exclusively or preferentially for bus traffic, including separated lanes. | Line Length Sum and Count |
| Bus Stops | Locations for passenger boarding/alighting on bus routes. | Buffer Count |
| Subway Lines | Length and number of rapid transit (subway) lines serving the area. | Line Length Sum and Count |
| Subway Stops | Transit access points on subway lines. | Buffer Count |
| Traffic Death and Serious Injuries | Pedestrians killed or seriously injured per mile of roadway (KSI) in the neighborhood tabulation area. | Buffer Mean |

| | | |
|---|---|---|
| Transportation and Noise Pollution | Percentage of tract area experiencing average annual noise above 45 db. | Buffer Mean |
| **Land Use** | | |
| Commercial Area | Land use representing commercial, retail, marketplace use (code=1). | Buffer and Land Use Intersection |
| Industrial Area | Land use representing industrial, warehouse, factory (code=2). | |
| Residential Area | Land use representing residential/apartment (code=3). | |
| Green Space Area | Parks, forest, grass, recreation grounds (code=4). | |
| Public Area | Institutional, educational, hospital, civic uses (code=5). | |
| Water Area | Water bodies, reservoirs, basins (code=6). | |
| Others | Other or unknown land use (code=0). | |

**Supplementary Table 2**. Binary Classification Performance Using Never Smoker Patients

| Model | Accuracy [a] | Macro-F1[b] | Weighted-F1[c] | ROC AUC [d] | Category-specific F1 Score | |
|---|---|---|---|---|---|---|
| Auto Gluon | | | | | | |
| Basic model | 0.58 | 0.49 | 0.53 | 0.672 | Negative | 0.28 |
| | | | | | Positive | 0.71 |
| Basic + urban factors | 0.59 | 0.44 | 0.48 | 0.677 | Negative | 0.14 |
| | | | | | Positive | 0.73 |
| Light GBM | | | | | | |
| Basic model | 0.62 | 0.49 | 0.53 | 0.712 | Negative | 0.23 |
| | | | | | Positive | 0.74 |
| Basic + urban factors | **0.63** | **0.50** | **0.54** | **0.685** | **Negative** | **0.24** |
| | | | | | **Positive** | **0.76** |

[a] **Accuracy:** The proportion of correctly classified observations out of the total number of observations, reflecting the overall correctness of the model.

[b] **Macro-F1:** The average F1 score (harmonic mean of precision and recall) calculated independently for each class. It treats all classes equally, making it sensitive to performance on minority classes.

[c] **Weighted-F1:** The average F1 score calculated for each class, weighted by the number of true instances (support) in each class. It accounts for class imbalance by giving more weight to the majority class.

[d] **ROC AUC:** The Area Under the Receiver Operating Characteristic Curve. It measures the model's ability to distinguish between classes across all classification thresholds (ranges from 0.5 for random guessing to 1.0 for perfect discrimination).

**Supplementary Table 3**. Binary Classification Performance Using WCM Data Only

| Model | Accuracy [a] | Macro-F1[b] | Weighted-F1[c] | ROC AUC [d] | Category-specific F1 Score | |
|---|---|---|---|---|---|---|
| Auto Gluon | | | | | | |
| Basic model | 0.71 | 0.67 | 0.72 | 0.732 | Negative | 0.79 |
| | | | | | Positive | 0.54 |
| Basic + urban factors | 0.68 | 0.64 | 0.69 | 0.732 | Negative | 0.76 |
| | | | | | Positive | 0.53 |
| Light GBM | | | | | | |
| Basic model | 0.75 | 0.70 | 0.76 | 0.764 | Negative | 0.82 |
| | | | | | Positive | 0.59 |
| Basic + urban factors | **0.72** | **0.68** | **0.73** | **0.755** | **Negative** | **0.80** |
| | | | | | **Positive** | **0.57** |

**Supplementary Table 4**. Multi-Classification Performance

| Model | Accuracy [a] | Macro-F1[b] | Weighted-F1[c] | ROC AUC [e] | Category-specific F1 Score | |
|---|---|---|---|---|---|---|
| Auto Gluon | | | | | | |
| Basic model | 0.57 | 0.51 | 0.57 | 0.692 | Negative | 0.50 |
| | | | | | Positive | 0.36 |
| | | | | | Unknown | 0.67 |
| Basic + urban factors | 0.63 | 0.55 | 0.62 | 0.791 | Negative | 0.54 |
| | | | | | Positive | 0.36 |
| | | | | | Unknown | 0.75 |
| Light GBM | | | | | | |
| Basic model | 0.57 | 0.50 | 0.55 | 0.698 | Negative | 0.48 |
| | | | | | Positive | 0.35 |
| | | | | | Unknown | 0.66 |
| Basic + urban factors | **0.66** | **0.59** | **0.65** | **0.806** | Negative | **0.58** |
| | | | | | Positive | **0.42** |
| | | | | | Unknown | **0.76** |

[e] **ROC AUC:** Weighted multiclass ROC AUC computed using a one-vs-rest strategy; per-class AUCs were averaged weighted by class prevalence. The average of per-class AUC values weighted by the class-specific proportion of samples, measuring the model’s discriminative ability while accounting for class imbalance by giving greater influence on more prevalent classes.

**Supplementary Figure 1.** Cohort Selection Diagram

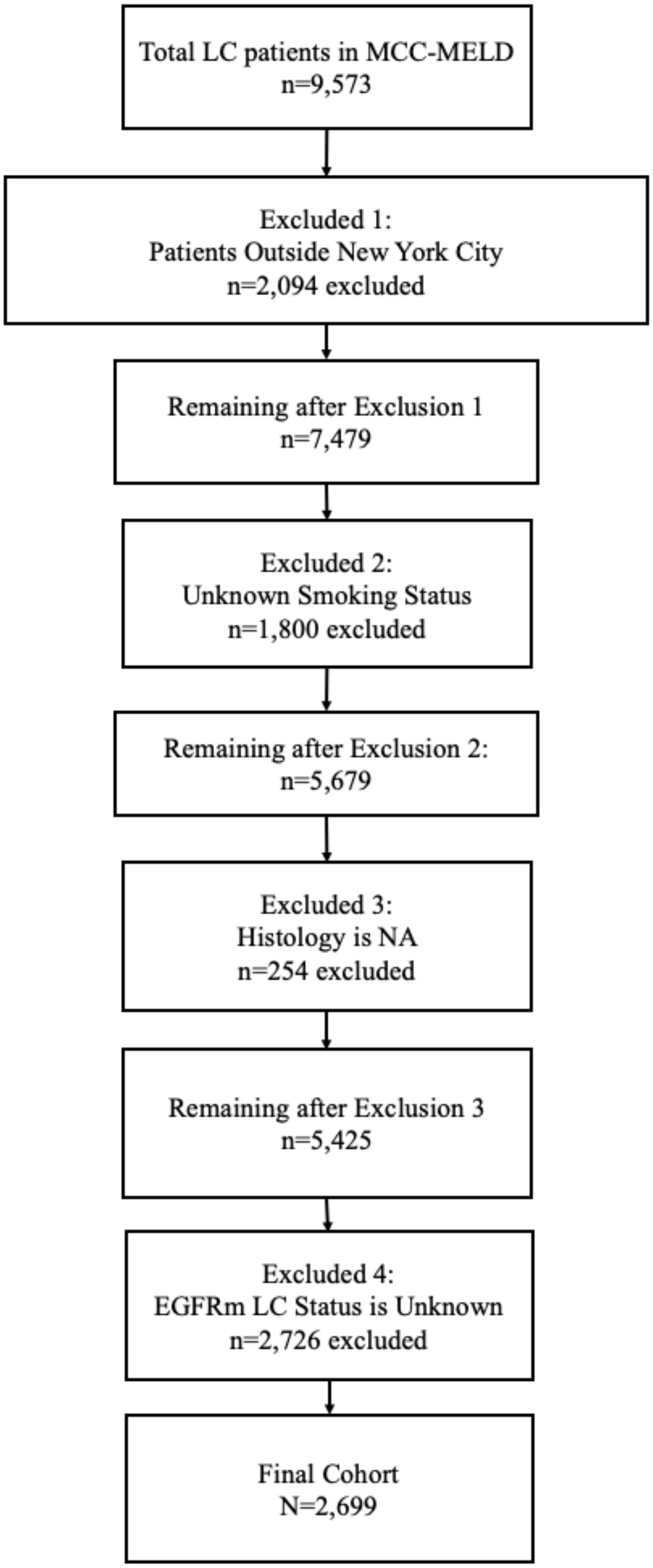

**Supplementary Figure 2.** Distribution of Lung Cancer Cases by Never or Ever Smoking Status and *EGFR*-mutant Status Across Meyer Cancer Center Catchment Areas at WCM (Manhattan), BMH (Brooklyn), and NYPQ (Queens).
Panels show: (a) *EGFR*-positive never smokers, (b) *EGFR*-negative never smokers, (c) *EGFR*-positive ever smokers, and (d) *EGFR*-negative ever smokers.

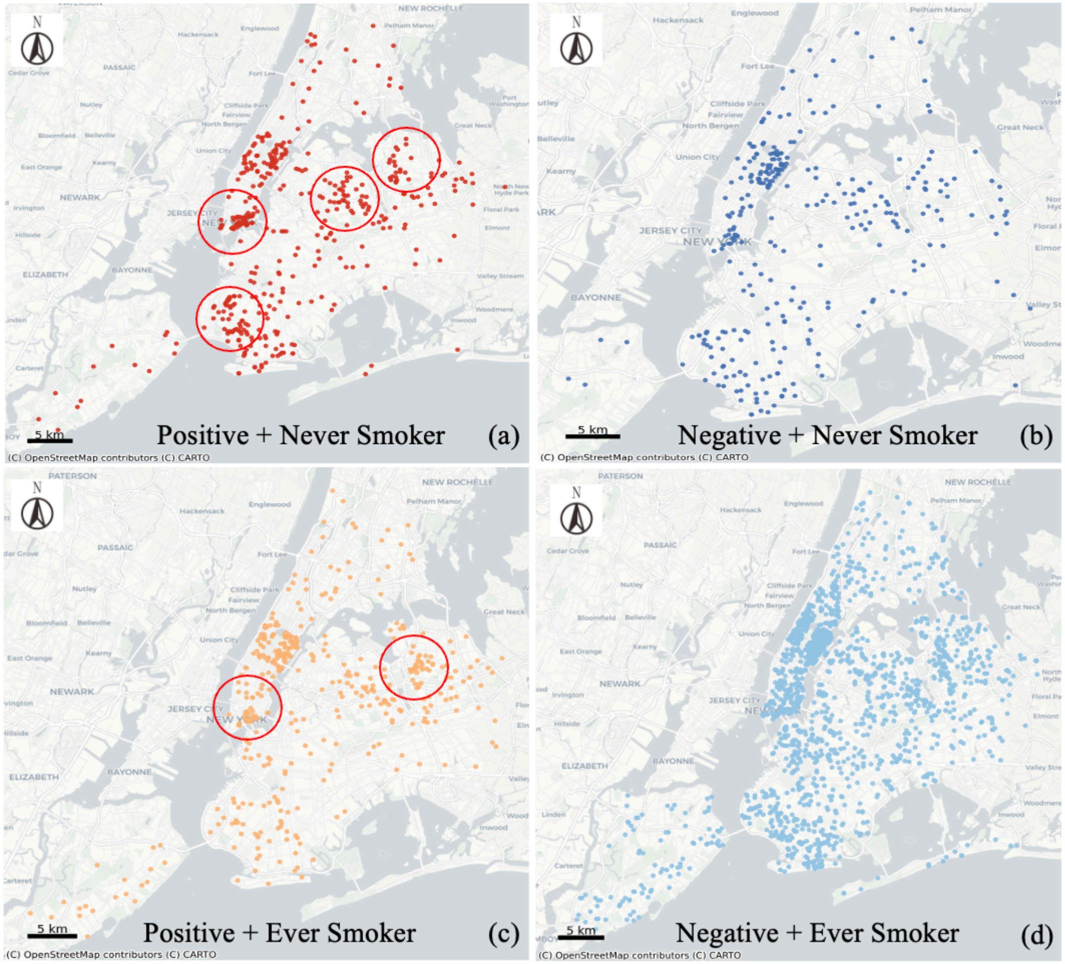

**Supplementary Figure 3**. Distribution of Lung Cancer Cases by Race and *EGFR*-mutant Status Across Meyer Cancer Center Catchment Areas at WCM (Manhattan), BMH (Brooklyn), and NYPQ (Queens).
Panels show: (a) *EGFR*-positive cases across different races and (b) *EGFR*-negative cases across different races.

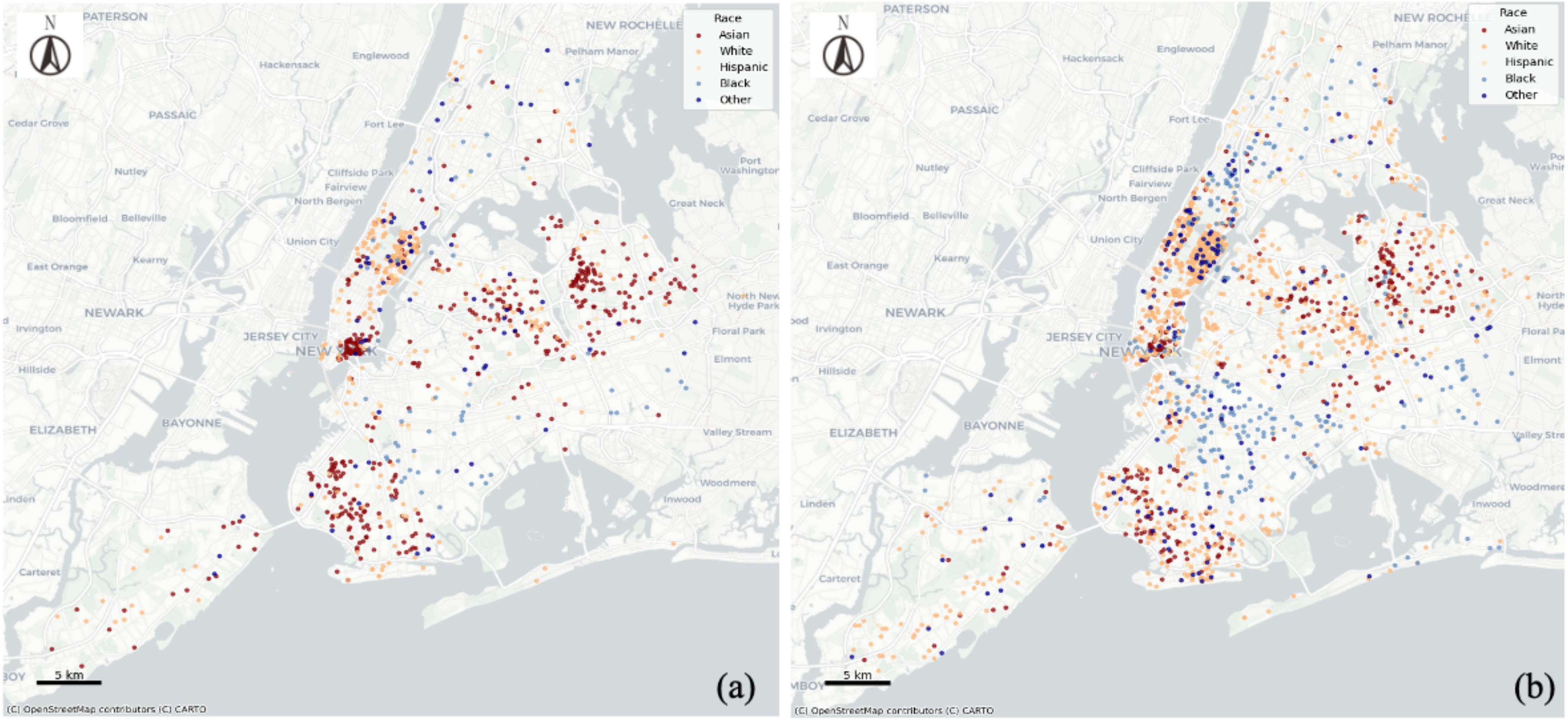

**Supplementary Figure 4**. Distribution of Lung Cancer Cases by Histology and *EGFR*-mutant Status Across Meyer Cancer Center Catchment Areas at WCM (Manhattan), BMH (Brooklyn), and NYPQ (Queens).
Panels show: (a) *EGFR*-positive cases across different histologic subtypes and (b) *EGFR*-negative cases across different histologic subtypes.

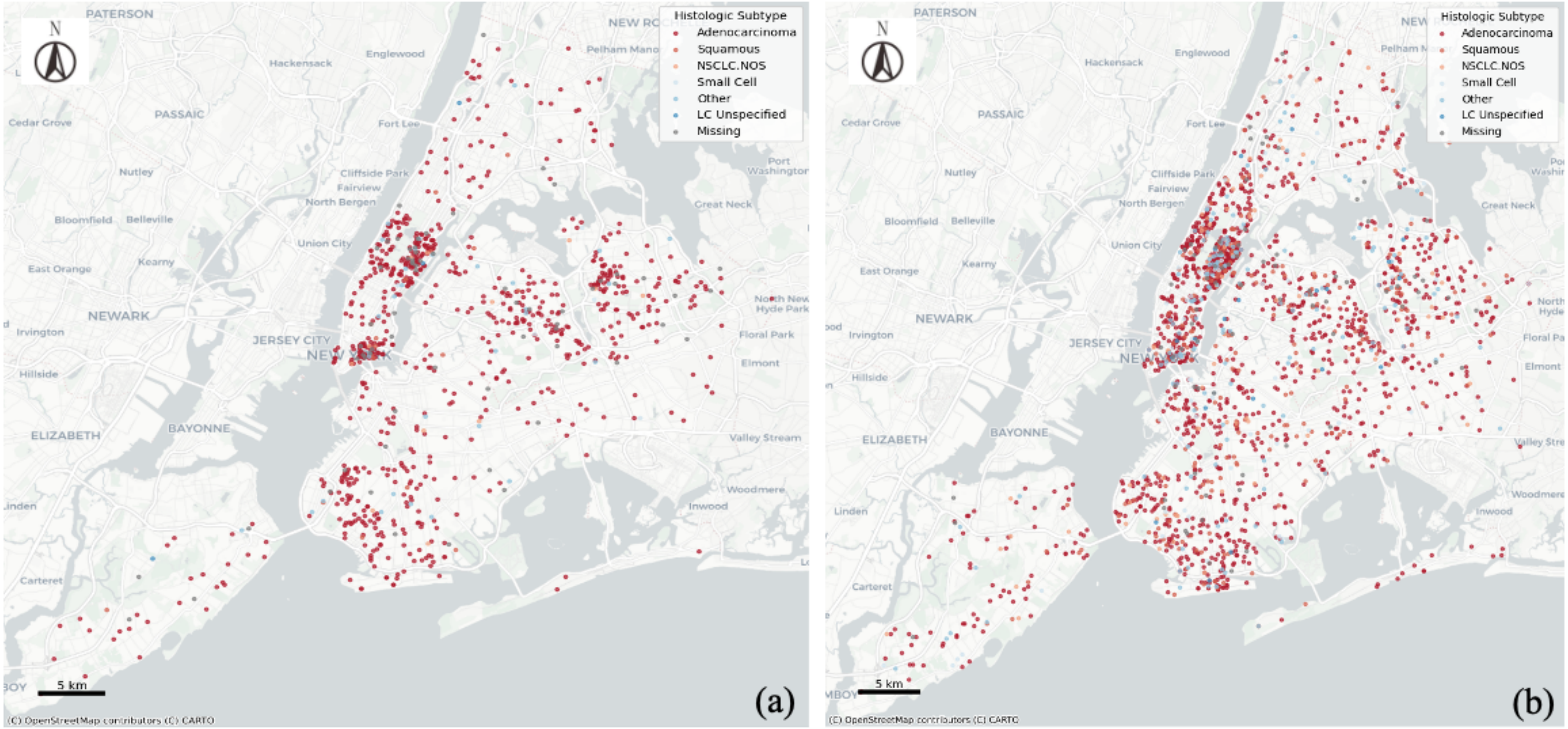

**Supplementary Figure 5.** Feature Importance of Model Using Never Smoker Patients

Abbreviations: SHAP, Shapley Additive exPlanations; hist_ix1, histology category; AD, adenocarcinoma; Noise_Abov, transportation and noise pollution above threshold; VMT, vehicle miles traveled; cum5_$NO_2$, cumulative 5-year nitrogen dioxide exposure; cum5_$PM_{2.5}$, fine particulate matter with aerodynamic diameter ≤ 2.5 µm.

Panels show: (a) Global feature importance, (b) SHAP summary plot

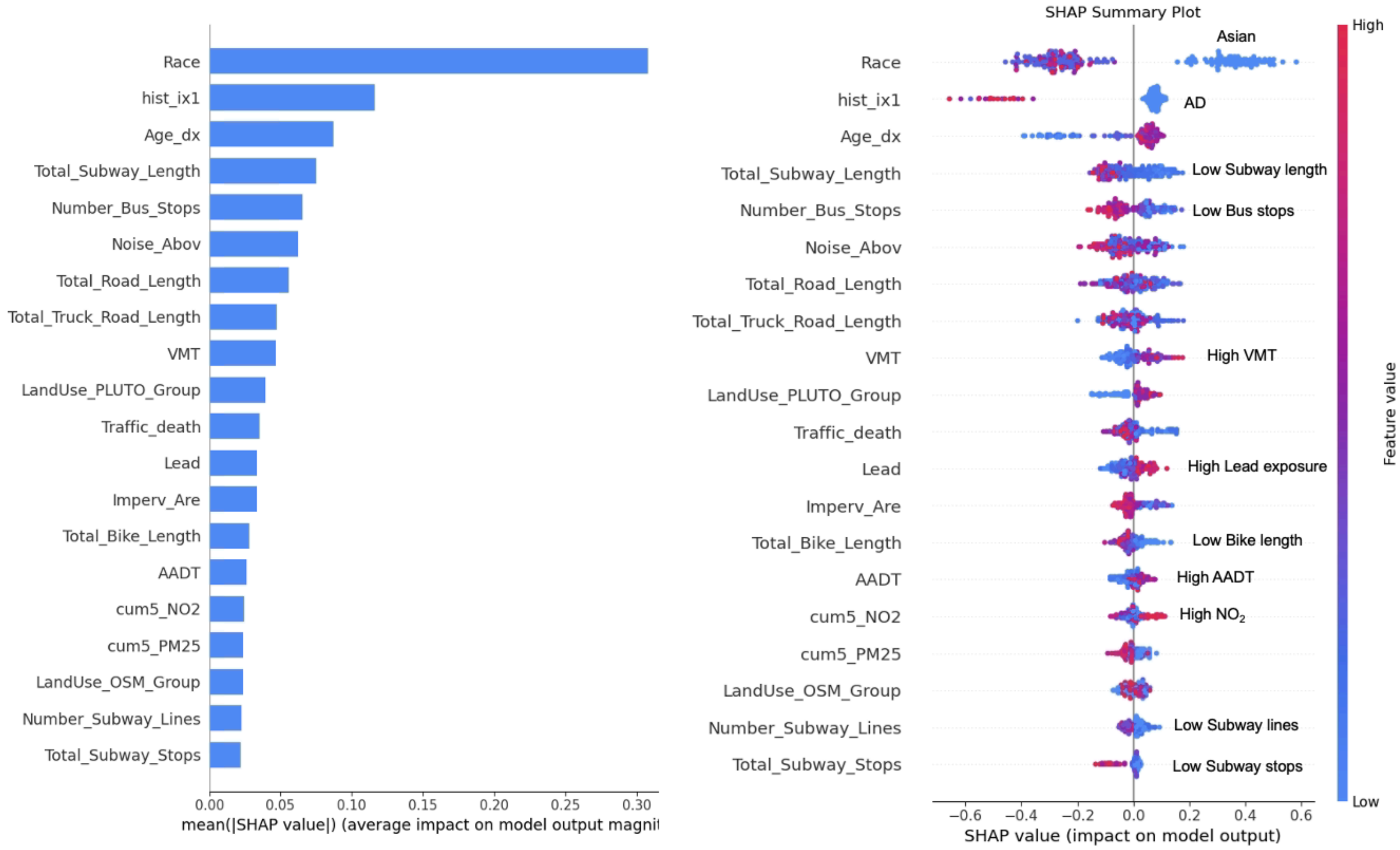


(a) (b)

**Supplementary Figure 6.** Feature Importance of Model Using WCM Data Only

Abbreviations: SHAP, Shapley Additive exPlanations; hist_ix1, histology category; AD, adenocarcinoma; Noise_Abov, transportation and noise pollution above threshold; VMT, vehicle miles traveled; cum5_$PM_{2.5}$, fine particulate matter with aerodynamic diameter ≤ 2.5 µm; cum5_BC, cumulative 5-year black carbon exposure; AADT, Annual average daily traffic; CISB, chemically intensive small business; homes_ac, air conditioning access.

Panels show: (a) Global feature importance, (b) SHAP summary plot

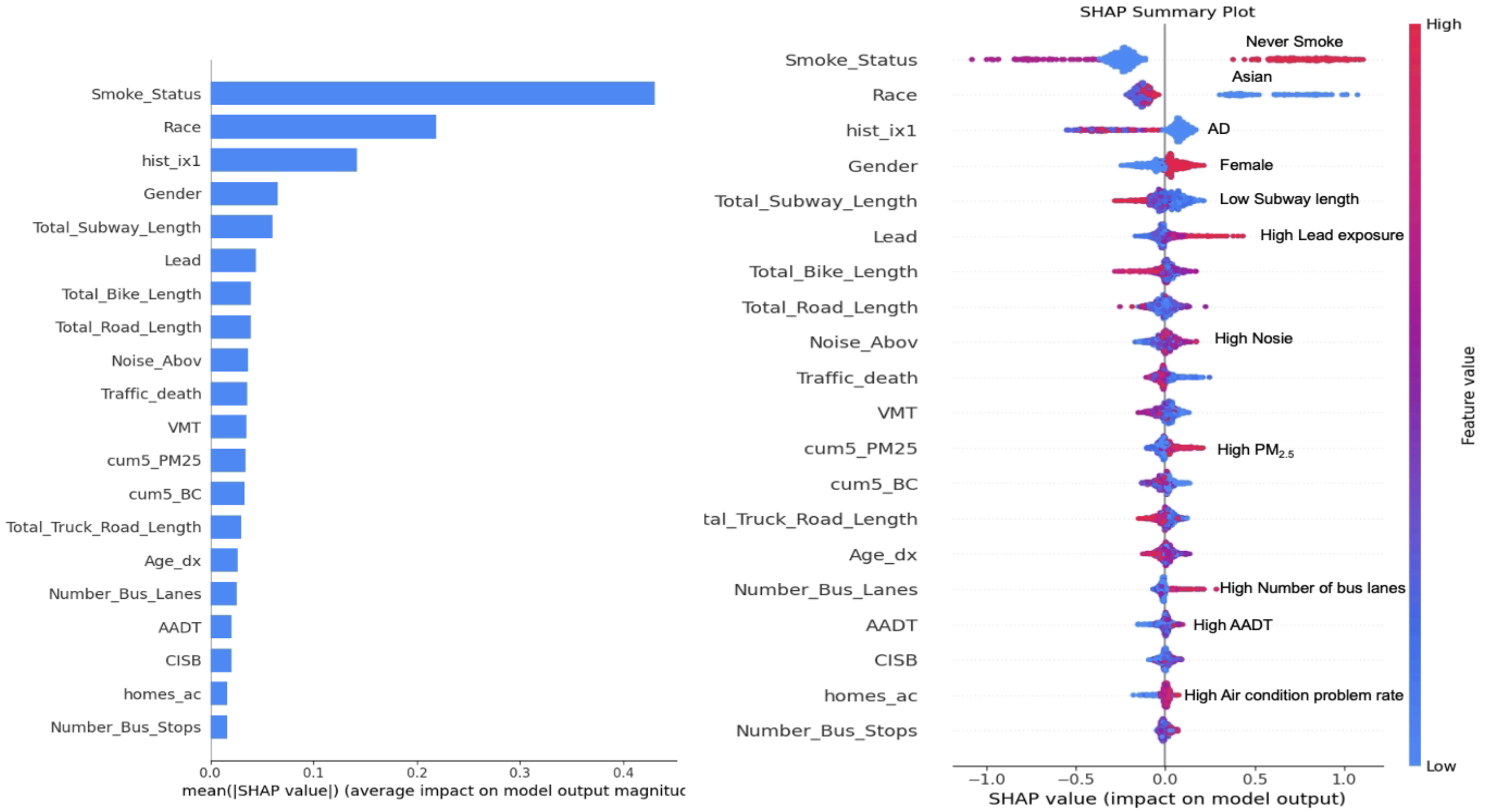


(a) (b)

**Supplementary Figure 7.** Feature Importance of Predictors Associated with *EGFR*-mutant Lung Cancer Using Light Gradient Boosting Machine (Light GBM)

Abbreviations: *EGFR*, epidermal growth factor receptor; SHAP, Shapley Additive exPlanations; hist_ix1, histology category; AD, adenocarcinoma; cum5_BC, cumulative 5-year black carbon exposure; cum5_$NO_2$, cumulative 5-year nitrogen dioxide exposure; cum5_$PM_{2.5}$, fine particulate matter with aerodynamic diameter ≤ 2.5 µm; lsl_pop_norm, housing maintenance issues.

Panels show: (a) Global feature importance, (b) *EGFR*-mutant lung cancer status is positive, (c) *EGFR*-mutant lung cancer status is unknown.

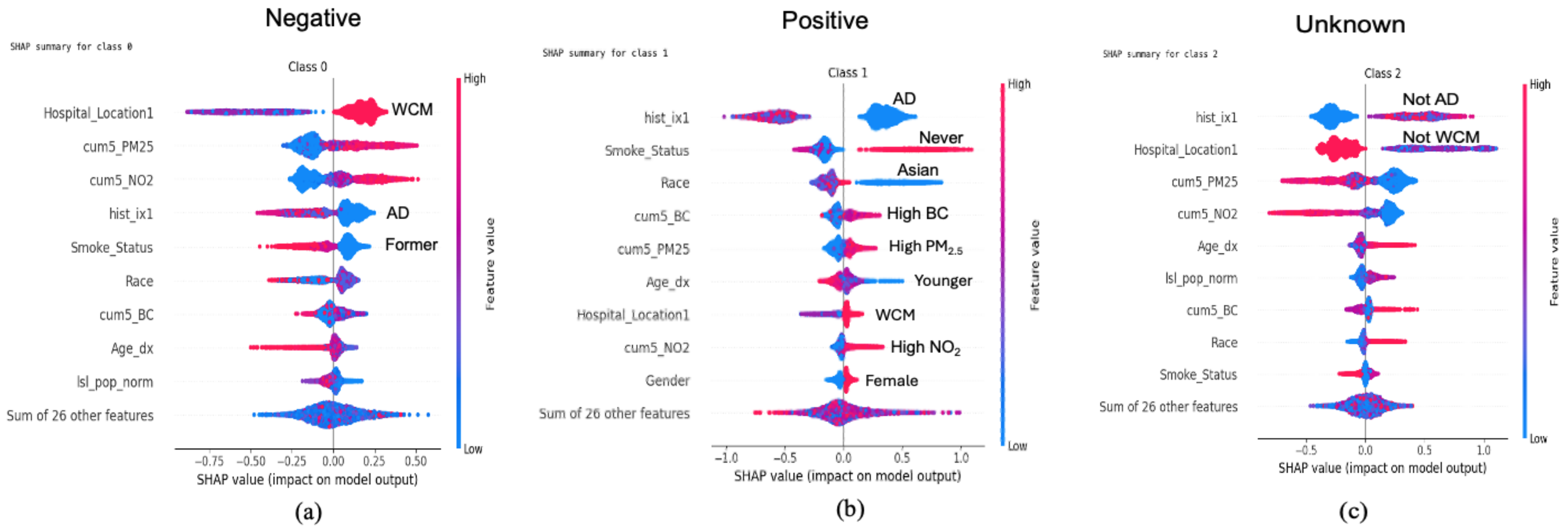

**Supplementary Methods 1.** Light Gradient Boosting Model (LightGBM)

LightGBM is a gradient boosting decision tree (GBDT) algorithm that iteratively builds additive decision trees to minimize a differentiable loss function. At boosting step $t$, the objective optimized is:

$$L^{(t)} = \sum_{i=1}^{n} l\left(y_i, \hat{y}_i^{(t-1)} + f_t(x_i)\right) + \Omega(f_t) \quad (1)$$

Where $f_t$ is the tree added at iteration $t$ and $\Omega(f_t)$ represents the regulation penalty. For computation efficiency, LightGBM applies a second-order Taylor expansion:

$$L^{(t)} \approx \sum_{i=1}^{n} \left(g_i f_t(x_i) + \frac{1}{2} h_i f_t(x_i)^2\right) + \Omega(f_t) \quad (2)$$

Where $g_i$ and $h_i$ denoting the first- and second-order gradients.

Tree splits are evaluated using the gain function:

$$Gain = \frac{1}{2}\left(\frac{G_L^2}{H_L+\lambda} + \frac{G_R^2}{H_R+\lambda} - \frac{(G_L+G_R)^2}{H_L+H_R+\lambda}\right) - \gamma \quad (3)$$

Where, $G_L$ , $G_R$ and $H_L, H_R$ are aggregated gradients for the potential child nodes. This formulation ensures efficient split selection while controlling tree complexity through λ and $\gamma$.

The LightGBM model was trained using a filtered subset of the dataset in which the binary target variable of *EGFR*-mutant LC status was restricted to classes 0 (negative) and 1 (positive). Predictor variables included demographic, environmental, transportation, and land-use characteristics. The data were split into training and testing subsets using a stratified 70/30 split to preserve class proportions. Additionally, model performance was evaluated using bootstrap resampling with 1,000 iterations, from which the 95% confidence intervals were derived.

In this dataset, the positive class was underrepresented, class imbalance was addressed using a weighting strategy based on:

$$scale\_pos\_weight = \frac{N_{train}}{N_{positive}} \quad (4)$$

This weighting increases the penalty for misclassifying minority-class samples, improving model sensitivity to rare outcomes during training.

These parameters including learning rate, number of leaves, feature fraction, bagging fraction, bagging frequency, and evaluation metrics were selected to balance predictive performance, computational efficiency, and regularization. The model was trained for up to 1,000 boosting iterations, with early stopping to prevent overfitting. Both training and validation sets were monitored, and training stopped automatically when no improvement occurred within 500 rounds. Each decision tree added to the ensemble minimized the second-order approximated loss. Formally, at each iteration:

$$f_t = arg\ arg\ L^{(t)} \quad (5)$$

After training, predicted probabilities were generated for the validation dataset. Instead of applying LightGBM's default threshold, the classification threshold was optimized to maximize the F1-score. The precision–recall curve was computed, and the optimal threshold was defined as:

$$\theta^* = arg\ arg\ F\ \ 1(\theta) \quad (6)$$

This optimized threshold was used to convert probability outputs into binary predictions for model evaluation. Model validation was performed using the held-out test set.
